# Development and 10-year validation of QRISK4-Lifetime for cardiovascular risk prediction in 13 million adults

**DOI:** 10.64898/2026.11.07.26367485

**Authors:** Julia Hippisley-Cox, Carol Coupland

**Affiliations:** Clinical Epidemiology & Predictive Medicine, Queen Mary University of London; Medical Statistics, Queen Mary University of London; Medical Statistics in Primary Care, University of Nottingham

## Abstract

**Objective:** To develop QRISK4-Lifetime to estimate cardiovascular disease (CVD) risk to age 95, compare its 10-year performance with QRISK4 and examine differences in risk classification between the two time horizons.

**Methods:** We used the linked electronic health datasets and cohort framework of the original QRISK4 study. We fitted sex-specific cause-specific Cox models for incident CVD and death from other causes, using attained age at cohort entry. Predicted risks combined both cause-specific hazards. We assessed discrimination and calibration at 10 years overall, by age and ethnicity. We compared characteristics of patients with a 10-year QRISK4-Lifetime risk ≥ 10% with an equal-sized group selected using lifetime risk to age 95

**Results:** There were 202,424 CVD events in the English derivation cohort of 9,976,306 people and 65,253 events in the validation cohort of 3,246,602 people. Overall C-indices were 0.860 (95% CI 0.858 to 0.862) in women and 0.845 (0.843 to 0.847) in men for both QRISK4-Lifetime and QRISK4. Observed risks were close to predicted risks. Each risk classification approach selected 310,648 patients (9.6%); 262,417 (8.1% of the cohort) met only the lifetime threshold. Of these, 72.8% were younger than 40 years and 79.5% were men.

**Conclusions:** QRISK4-Lifetime had similar performance to QRISK4 when evaluated at 10 years. Lifetime assessment identified a substantially younger population with increased lifetime risks but below the specified 10-year threshold of 10%, supporting its potential complementary role in prevention discussions. The validity of lifetime estimates and the clinical consequences of using both time horizons require further evaluation.

**Key messages:** *What is already known on this topic:* Age strongly influences ten-year cardiovascular risk, which can be low in younger adults with adverse risk factors. CVD lifetime estimates can support earlier discussions about smoking, other modifiable risk factors and prevention among people whose short-term risk remains low.

*What this study adds:* QRISK4-Lifetime had similar 10-year discrimination and calibration to QRISK4. QRISK4-Lifetime enabled identification of a substantially younger high-risk group with a different ethnic and socioeconomic profile. QRISK4-Lifetime provides an integrated framework for estimating CVD risk over two time-horizons using the expanded QRISK4 predictor set within one tool.

*How this study might affect research practice or policy:* QRISK4-Lifetime identification of a substantially younger high-risk group with a different ethnic and socioeconomic profile below the specified short-term threshold supports its potential role in preventive risk communication. Further research is needed to evaluate its clinical utility and cost-effectiveness and how best to communicate results.

## Introduction

International guidelines from the United States of America,^1^ Canada,^2^ Europe^3^ and the United Kingdom^4^ recommend use of cardiovascular disease (CVD) risk prediction tools to identify people at high-risk who may benefit from preventive interventions. Risk assessment has traditionally focused on the probability of developing CVD within 10 years. However, because age is a major determinant of short-term CVD risk, younger adults with adverse risk factors may have low 10-year risk despite substantial risk over their remaining lifetime. Estimation of lifetime CVD risk may therefore complement short-term risk assessment by supporting discussions about prevention with individuals^5^ and informing projections of population CVD burden.^6,7^

UK guidance has recommended QRISK tools for CVD assessment since 2010^4^. In 2024, the fourth version (QRISK4) extended the currently recommended QRISK3 algorithm by incorporating additional clinical conditions and accounting for competing mortality.^8,9^ Here we develop QRISK4-Lifetime using the same datasets to estimate CVD risk over the remaining lifetime.^8^ We investigate whether lifetime assessment identifies different patients as high risk. We also compare its 10-year predictions with QRISK4 with particular attention to performance across ethnic groups, given recognised evidence gaps^3^ and the importance of equitable performance of CVD risk prediction models across population groups^3 10^.

## Methods

We used the linked datasets described in the original QRISK4 study.^8^ QResearch practices in England were randomly allocated to derivation (three-quarters) or validation (one-quarter) cohorts. The cohorts included adults aged 18–84 years between 1 January 2010 and 31 December 2021 without pre-existing CVD or statin treatment. Participants with missing Townsend scores were excluded. The derivation and validation cohorts included 9,976,306 and 3,246,602 participants, respectively. ^8^

Cohort entry was the latest of the study start (1^st^ January 2010), a participant’s 18^th^ birthday, and the practice registration date. The outcome was first recorded CVD (myocardial infarction, ischaemic heart disease, stroke or transient ischaemic attack) with death from other causes as the competing event. Outcomes were identified from primary care records, linked hospital and mortality records. Cohort exit was the earliest of first CVD event, death, deregistration from the practice, the participant’s 95^th^ birthday or study end (31 December 2021).

Separate cause-specific Cox models were fitted for incident CVD and non-CVD death in women and men, using age as the underlying time scale and accounting for delayed entry (left truncation) using age at cohort entry.^11^ We combined the CVD and competing-risk non-CVD death hazards to estimate cumulative incidence.^12^ This approach allows estimation of risk predictions over short and longer risk horizons.^5,11,12^ We evaluated 10-year risk and lifetime risk defined as the probability of a first CVD event before age 95, conditional on being alive and CVD-free at the assessment age.^5 12^ Predictor definitions and handling of missing data followed the original study.^8^ QRISK4 included established demographic and clinical predictors in QRISK3, together with new predictors for brain, lung, blood and oral cancers, chronic obstructive pulmonary disease (COPD), learning disability and Down’s syndrome and pre-eclampsia and postnatal depression in women. Interactions between predictors and fractional polynomial terms for age at cohort entry followed the specification in the original QRISK4 study, allowing predictor associations to vary with baseline age.^8^ Model development used five imputed datasets separately by sex, with coefficient estimates combined using Rubin’s rules.^13^

In the validation cohort, we compared the performance of QRISK4-Lifetime with QRISK4 both using a 10-year horizon. We estimated C-indices overall and by age and ethnicity accounting for competing risks.^14^ Calibration was assessed using observed cumulative incidence, centile plots, intercepts and slopes based on risk-stratified pseudovalues accounting for competing risks.^15^

We compared participant characteristics in the validation cohort classified as high-risk based on a QRISK4-Lifetime 10-year risk ≥10% with an equal-sized high-risk group based on QRISK4-Lifetime predicted lifetime risk. We estimated observed 10-year CVD cumulative incidence (95% CI), overall and by age, sex and ethnicity.

All eligible participants were included. Stata v18 was used for analyses.

## Results

There were 202,424 incident CVD events in the derivation cohort of 9,976,306 people during 49.1 million person-years and 65,253 incident CVD events in the validation cohort of 3,246,602 people during 16.0 million person years.^8^ The adjusted hazard ratios in the QRISK4-Lifetime CVD models were very similar to those in the QRISK4 models. ^8,11^

Table 1 shows the same 10-year discrimination for both QRISK4 and QRISK4-lifetime in the validation cohort. 10-year C-index values for QRISK4-lifetime and QRISK4 were 0.860 (0.858 to 0.862) in women and 0.845 (0.843 to 0.847) in men. Discrimination values were similar between models across age and ethnic groups and tended to be higher in younger ages. Calibration slopes, intercepts and plots show similar calibration for QRISK4 and QRISK4-Lifetime at 10 years (Table 1 and Figure 1).

**Table 1.** Discrimination and risk stratified pseudovalue calibration slope and intercept for QRISK4 and QRISK4-Lifetime evaluated at 10 years in the QResearch validation cohort overall, by age at cohort entry and ethnic group. Values are estimates with 95% confidence intervals.

|  |  | **Women** |  | **Men** |  |
|---|---|---|---|---|---|
|  |  | QRISK4-10-year estimate (95% CI) | QRISK4-lifetime at 10 years estimate (95% CI) | QRISK4-10-year estimate (95% CI) | QRISK4-lifetime at 10 years estimate (95% CI) |
| ***C-Index*** | ***Overall*** | **0.860 (0.858 to 0.862)** | **0.860 (0.858 to 0.862)** | **0.845 (0.843 to 0.847)** | **0.845 (0.843 to 0.847)** |
| Age | <45 | 0.779 (0.768 to 0.791) | 0.779 (0.767 to 0.790) | 0.807 (0.800 to 0.813) | 0.807 (0.800 to 0.813) |
|  | 45-64 | 0.710 (0.704 to 0.715) | 0.710 (0.705 to 0.716) | 0.684 (0.680 to 0.688) | 0.684 (0.680 to 0.688) |
|  | 65+ | 0.651 (0.646 to 0.656) | 0.651 (0.646 to 0.655) | 0.609 (0.604 to 0.615) | 0.610 (0.604 to 0.615) |
| Ethnic Group | White | 0.850 (0.848 to 0.852) | 0.850 (0.848 to 0.853) | 0.832 (0.830 to 0.834) | 0.832 (0.830 to 0.834) |
|  | Indian | 0.881 (0.868 to 0.895) | 0.881 (0.868 to 0.895) | 0.868 (0.858 to 0.878) | 0.868 (0.858 to 0.878) |
|  | Pakistani | 0.856 (0.836 to 0.876) | 0.855 (0.834 to 0.875) | 0.865 (0.853 to 0.877) | 0.865 (0.853 to 0.877) |
|  | Bangladeshi | 0.879 (0.856 to 0.902) | 0.879 (0.857 to 0.902) | 0.855 (0.842 to 0.869) | 0.856 (0.843 to 0.870) |
|  | Other Asian | 0.891 (0.875 to 0.906) | 0.890 (0.874 to 0.906) | 0.853 (0.838 to 0.868) | 0.853 (0.838 to 0.868) |
|  | Caribbean | 0.837 (0.822 to 0.852) | 0.837 (0.822 to 0.852) | 0.820 (0.805 to 0.835) | 0.820 (0.804 to 0.835) |
|  | Black African | 0.866 (0.849 to 0.884) | 0.866 (0.848 to 0.884) | 0.854 (0.840 to 0.867) | 0.853 (0.840 to 0.866) |
|  | Chinese | 0.891 (0.825 to 0.956) | 0.892 (0.832 to 0.953) | 0.918 (0.902 to 0.935) | 0.918 (0.901 to 0.934) |
|  | Other group | 0.870 (0.854 to 0.885) | 0.870 (0.856 to 0.885) | 0.869 (0.859 to 0.879) | 0.868 (0.858 to 0.878) |
| ***Slope<sup>\*</sup>*** | ***Overall*** | **1.001 (0.994 to 1.007)** | **0.994 (0.987 to 1.000)** | **1.002 (0.997 to 1.008)** | **1.002 (0.996 to 1.008)** |
| Age | <45 | 0.992 (0.979 to 1.005) | 0.983 (0.970 to 0.996) | 1.006 (0.995 to 1.016) | 0.999 (0.988 to 1.009) |
|  | 45-64 | 1.005 (0.997 to 1.014) | 1.001 (0.993 to 1.009) | 0.996 (0.988 to 1.003) | 1.004 (0.997 to 1.011) |
|  | 65+ | 0.999 (0.988 to 1.009) | 0.990 (0.979 to 1.000) | 1.015 (1.002 to 1.028) | 1.000 (0.987 to 1.012) |
| Ethnic Group | White | 0.998 (0.991 to 1.005) | 0.988 (0.981 to 0.995) | 1.004 (0.997 to 1.010) | 1.000 (0.993 to 1.006) |
|  | Indian | 1.038 (0.994 to 1.082) | 1.052 (1.008 to 1.096) | 1.011 (0.979 to 1.043) | 1.019 (0.986 to 1.051) |
|  | Pakistani | 0.974 (0.924 to 1.024) | 1.004 (0.952 to 1.057) | 0.966 (0.927 to 1.005) | 1.002 (0.961 to 1.044) |
|  | Bangladeshi | 1.001 (0.937 to 1.065) | 1.018 (0.953 to 1.084) | 1.014 (0.968 to 1.060) | 1.066 (1.016 to 1.116) |
|  | Other Asian | 1.017 (0.964 to 1.070) | 1.024 (0.972 to 1.077) | 1.033 (0.989 to 1.076) | 1.040 (0.996 to 1.084) |
|  | Caribbean | 1.031 (0.986 to 1.077) | 1.050 (1.004 to 1.097) | 0.983 (0.943 to 1.023) | 1.014 (0.972 to 1.056) |
|  | Black African | 0.975 (0.934 to 1.015) | 1.006 (0.964 to 1.048) | 0.977 (0.943 to 1.011) | 1.011 (0.976 to 1.046) |
|  | Chinese | 1.041 (0.930 to 1.152) | 0.993 (0.891 to 1.094) | 1.006 (0.928 to 1.084) | 0.939 (0.870 to 1.009) |
|  | Other group | 1.030 (0.993 to 1.068) | 1.029 (0.992 to 1.066) | 0.987 (0.957 to 1.016) | 0.986 (0.957 to 1.015) |
| ***Intercept<sup>±</sup>*** | ***Overall*** | **-0.011 (-0.026 to 0.004)** | **0.005 (-0.010 to 0.021)** | **-0.017 (-0.030 to -0.005)** | **-0.016 (-0.028 to -0.003)** |
| Age | <45 | 0.025 (-0.037 to 0.088) | 0.051 (-0.012 to 0.113) | -0.014 (-0.055 to 0.026) | 0.003 (-0.038 to 0.044) |
|  | 45-64 | -0.034 (-0.059 to -0.008) | -0.013 (-0.038 to 0.011) | -0.006 (-0.023 to 0.011) | -0.022 (-0.039 to -0.005) |
|  | 65+ | -0.005 (-0.025 to 0.014) | 0.010 (-0.010 to 0.029) | -0.029 (-0.048 to -0.010) | -0.013 (-0.032 to 0.006) |
| Ethnic Group | White | -0.005 (-0.022 to 0.011) | 0.017 (0.001 to 0.034) | -0.017 (-0.030 to -0.004) | -0.010 (-0.023 to 0.004) |
|  | Indian | -0.088 (-0.194 to 0.018) | -0.110 (-0.215 to -0.006) | -0.037 (-0.111 to 0.036) | -0.048 (-0.121 to 0.025) |
|  | Pakistani | 0.000 (-0.130 to 0.130) | -0.054 (-0.184 to 0.076) | 0.026 (-0.063 to 0.115) | -0.041 (-0.129 to 0.048) |
|  | Bangladeshi | -0.035 (-0.215 to 0.146) | -0.076 (-0.255 to 0.103) | -0.128 (-0.241 to -0.014) | -0.224 (-0.336 to -0.112) |
|  | Other Asian | -0.069 (-0.210 to 0.073) | -0.079 (-0.218 to 0.059) | -0.092 (-0.193 to 0.009) | -0.099 (-0.200 to 0.002) |
|  | Caribbean | -0.093 (-0.202 to 0.016) | -0.127 (-0.235 to -0.019) | 0.018 (-0.079 to 0.115) | -0.045 (-0.142 to 0.051) |
|  | Black African | 0.070 (-0.056 to 0.195) | -0.011 (-0.134 to 0.112) | 0.071 (-0.024 to 0.167) | -0.011 (-0.106 to 0.083) |
|  | Chinese | -0.132 (-0.449 to 0.185) | 0.003 (-0.316 to 0.322) | -0.016 (-0.220 to 0.189) | 0.165 (-0.039 to 0.369) |
|  | Other group | -0.068 (-0.170 to 0.035) | -0.062 (-0.164 to 0.039) | 0.025 (-0.053 to 0.102) | 0.028 (-0.050 to 0.105) |

\*Slope: intercept fixed at zero; ideal value one. †Intercept: slope fixed at one; ideal value zero; positive values indicate underprediction.

**Figure 1:**
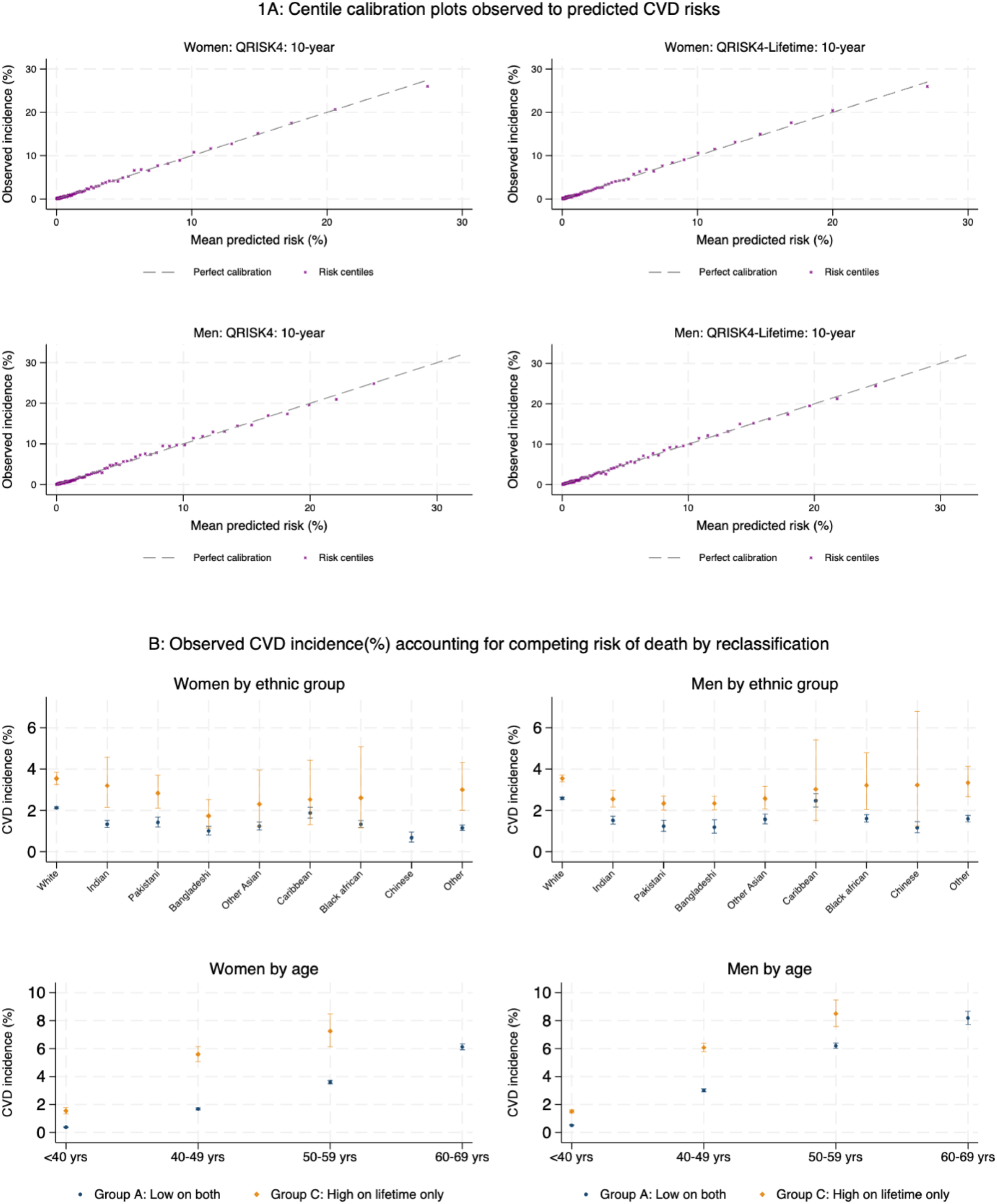
(A) Calibration of the observed and predicted CVD risks (B) observed cumulative CVD 10-year incidence (95%CI) by risk group, age, sex and ethnicity. Groups are defined in Table 2.

Table 2 compares characteristics of participants by reclassification thresholds for 10-year QRISK4-Lifetime and QRISK4-Lifetime to age 95, each identifying a total of 310,648 high-risk patients (9.6% of the validation cohort, giving a threshold of 59.3% for lifetime risk). Compared with Group A (low risk on both measures), Group C (high on lifetime risk but low on 10-year risk) included more participants who were under 40 years (72.8% vs 64.2%), more men (79.5% vs 43.2%), more in Indian, Pakistani, Bangladeshi and other Asian groups (total 32.9% vs 11.3%), more current smokers (44.4% vs 19.1%) and more with treated hypertension (6.0% vs 1.6%), migraine (15.0% vs 5.5%), erectile dysfunction (2.9% vs 1.0%) and pre-eclampsia (0.4% vs 0.2%).

**Table 2.**
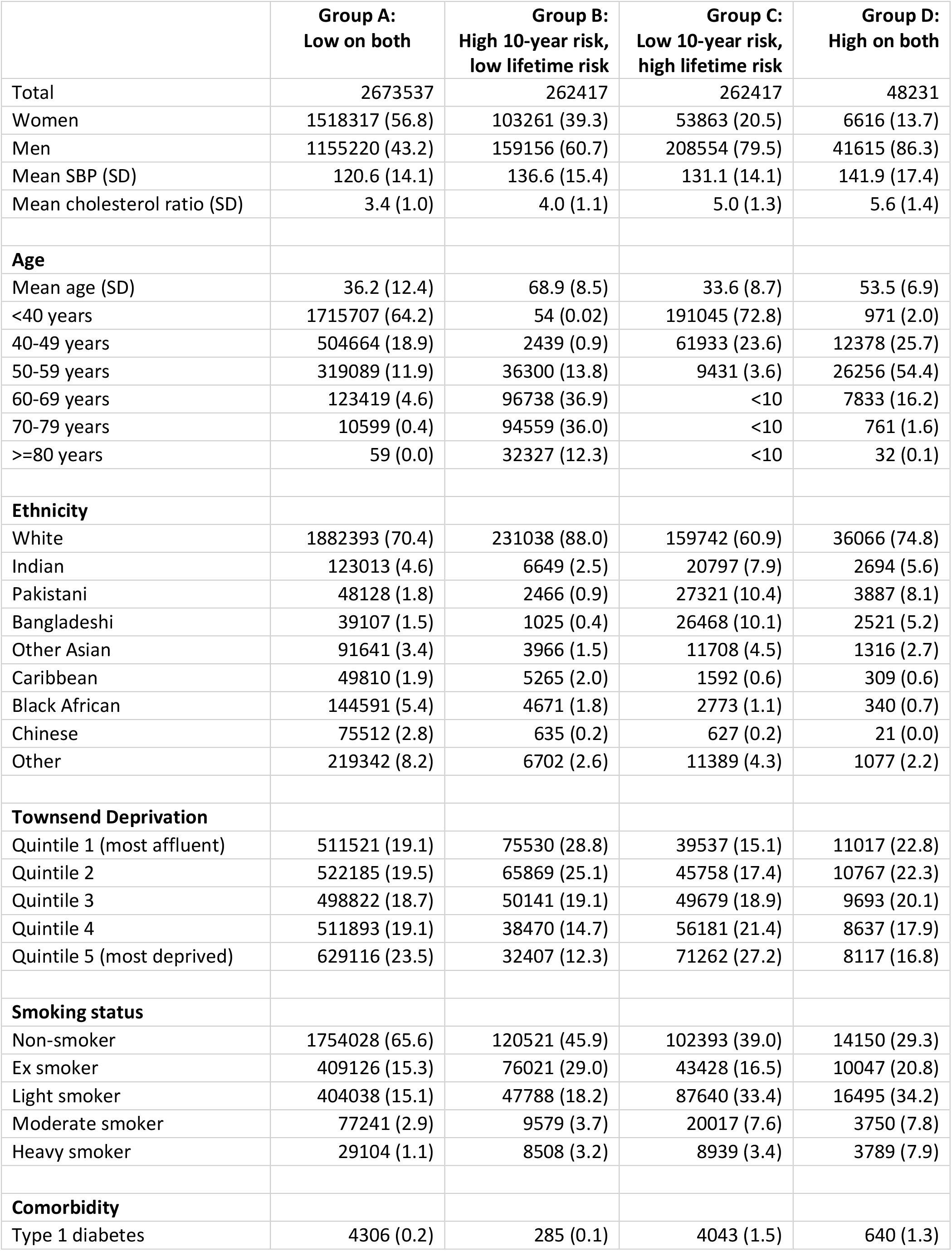
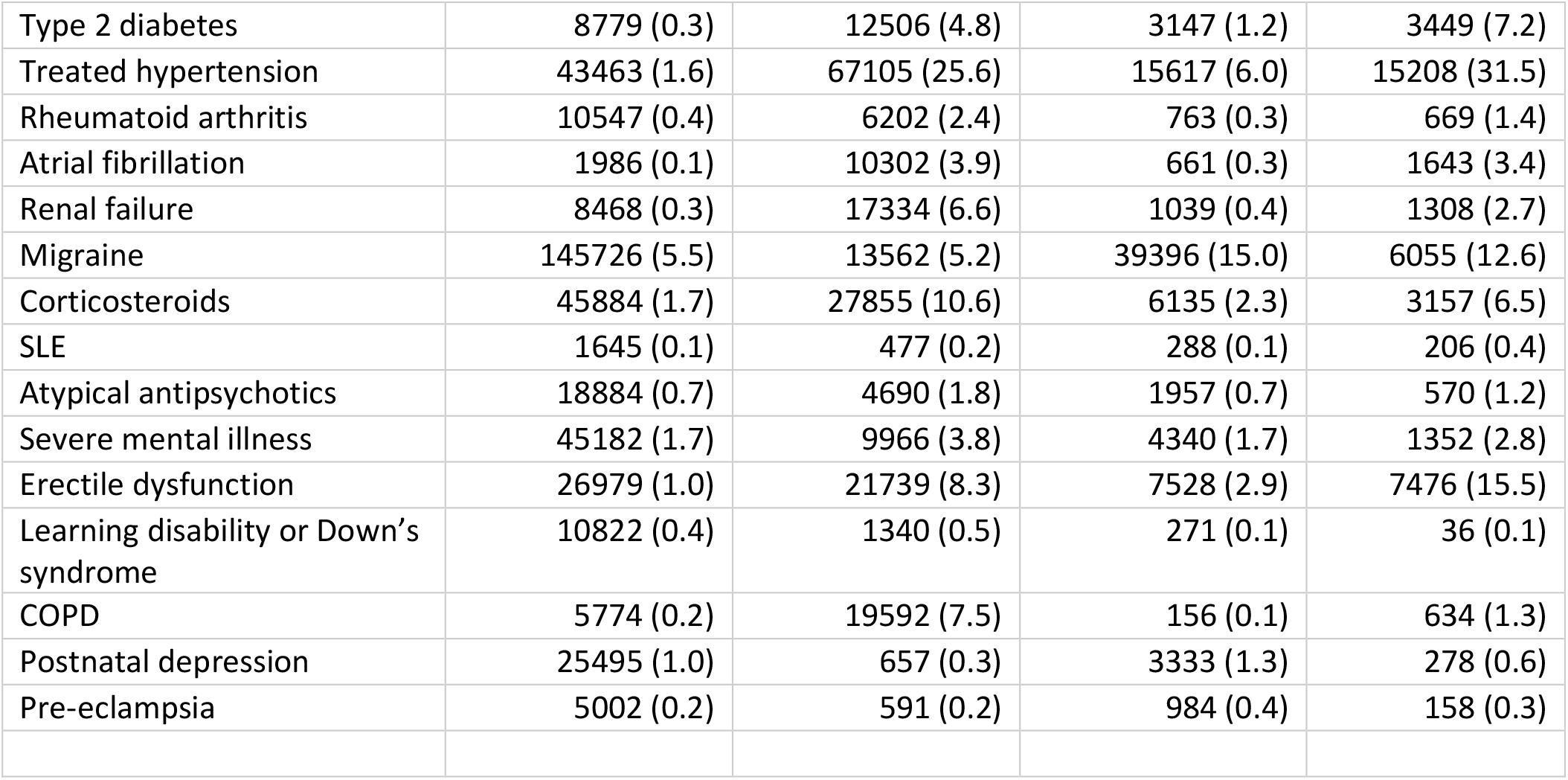
Risk group re-classification: Group A: Low on both QRISK4-lifetime (<59.3%) and 10-year risk (<10%); Group B: High on 10-year risk, low on lifetime risk; Group C: Low on 10-year risk high on lifetime risk; Group D: high on both lifetime and 10-year risk. Figures are counts and column percentages unless otherwise specified.

|  | Group A:<br>Low on both | Group B:<br>High 10-year risk, low lifetime risk | Group C:<br>Low 10-year risk, high lifetime risk | Group D:<br>High on both |
|---|---|---|---|---|
| Total | 2673537 | 262417 | 262417 | 48231 |
| Women | 1518317 (56.8) | 103261 (39.3) | 53863 (20.5) | 6616 (13.7) |
| Men | 1155220 (43.2) | 159156 (60.7) | 208554 (79.5) | 41615 (86.3) |
| Mean SBP (SD) | 120.6 (14.1) | 136.6 (15.4) | 131.1 (14.1) | 141.9 (17.4) |
| Mean cholesterol ratio (SD) | 3.4 (1.0) | 4.0 (1.1) | 5.0 (1.3) | 5.6 (1.4) |
| **Age** |  |  |  |  |
| Mean age (SD) | 36.2 (12.4) | 68.9 (8.5) | 33.6 (8.7) | 53.5 (6.9) |
| <40 years | 1715707 (64.2) | 54 (0.02) | 191045 (72.8) | 971 (2.0) |
| 40-49 years | 504664 (18.9) | 2439 (0.9) | 61933 (23.6) | 12378 (25.7) |
| 50-59 years | 319089 (11.9) | 36300 (13.8) | 9431 (3.6) | 26256 (54.4) |
| 60-69 years | 123419 (4.6) | 96738 (36.9) | <10 | 7833 (16.2) |
| 70-79 years | 10599 (0.4) | 94559 (36.0) | <10 | 761 (1.6) |
| >=80 years | 59 (0.0) | 32327 (12.3) | <10 | 32 (0.1) |
| **Ethnicity** |  |  |  |  |
| White | 1882393 (70.4) | 231038 (88.0) | 159742 (60.9) | 36066 (74.8) |
| Indian | 123013 (4.6) | 6649 (2.5) | 20797 (7.9) | 2694 (5.6) |
| Pakistani | 48128 (1.8) | 2466 (0.9) | 27321 (10.4) | 3887 (8.1) |
| Bangladeshi | 39107 (1.5) | 1025 (0.4) | 26468 (10.1) | 2521 (5.2) |
| Other Asian | 91641 (3.4) | 3966 (1.5) | 11708 (4.5) | 1316 (2.7) |
| Caribbean | 49810 (1.9) | 5265 (2.0) | 1592 (0.6) | 309 (0.6) |
| Black African | 144591 (5.4) | 4671 (1.8) | 2773 (1.1) | 340 (0.7) |
| Chinese | 75512 (2.8) | 635 (0.2) | 627 (0.2) | 21 (0.0) |
| Other | 219342 (8.2) | 6702 (2.6) | 11389 (4.3) | 1077 (2.2) |
| **Townsend Deprivation** |  |  |  |  |
| Quintile 1 (most affluent) | 511521 (19.1) | 75530 (28.8) | 39537 (15.1) | 11017 (22.8) |
| Quintile 2 | 522185 (19.5) | 65869 (25.1) | 45758 (17.4) | 10767 (22.3) |
| Quintile 3 | 498822 (18.7) | 50141 (19.1) | 49679 (18.9) | 9693 (20.1) |
| Quintile 4 | 511893 (19.1) | 38470 (14.7) | 56181 (21.4) | 8637 (17.9) |
| Quintile 5 (most deprived) | 629116 (23.5) | 32407 (12.3) | 71262 (27.2) | 8117 (16.8) |
| **Smoking status** |  |  |  |  |
| Non-smoker | 1754028 (65.6) | 120521 (45.9) | 102393 (39.0) | 14150 (29.3) |
| Ex smoker | 409126 (15.3) | 76021 (29.0) | 43428 (16.5) | 10047 (20.8) |
| Light smoker | 404038 (15.1) | 47788 (18.2) | 87640 (33.4) | 16495 (34.2) |
| Moderate smoker | 77241 (2.9) | 9579 (3.7) | 20017 (7.6) | 3750 (7.8) |
| Heavy smoker | 29104 (1.1) | 8508 (3.2) | 8939 (3.4) | 3789 (7.9) |
| **Comorbidity** |  |  |  |  |
| Type 1 diabetes | 4306 (0.2) | 285 (0.1) | 4043 (1.5) | 640 (1.3) |
| Type 2 diabetes | 8779 (0.3) | 12506 (4.8) | 3147 (1.2) | 3449 (7.2) |
| Treated hypertension | 43463 (1.6) | 67105 (25.6) | 15617 (6.0) | 15208 (31.5) |
| Rheumatoid arthritis | 10547 (0.4) | 6202 (2.4) | 763 (0.3) | 669 (1.4) |
| Atrial fibrillation | 1986 (0.1) | 10302 (3.9) | 661 (0.3) | 1643 (3.4) |
| Renal failure | 8468 (0.3) | 17334 (6.6) | 1039 (0.4) | 1308 (2.7) |
| Migraine | 145726 (5.5) | 13562 (5.2) | 39396 (15.0) | 6055 (12.6) |
| Corticosteroids | 45884 (1.7) | 27855 (10.6) | 6135 (2.3) | 3157 (6.5) |
| SLE | 1645 (0.1) | 477 (0.2) | 288 (0.1) | 206 (0.4) |
| Atypical antipsychotics | 18884 (0.7) | 4690 (1.8) | 1957 (0.7) | 570 (1.2) |
| Severe mental illness | 45182 (1.7) | 9966 (3.8) | 4340 (1.7) | 1352 (2.8) |
| Erectile dysfunction | 26979 (1.0) | 21739 (8.3) | 7528 (2.9) | 7476 (15.5) |
| Learning disability or Down's syndrome | 10822 (0.4) | 1340 (0.5) | 271 (0.1) | 36 (0.1) |
| COPD | 5774 (0.2) | 19592 (7.5) | 156 (0.1) | 634 (1.3) |
| Postnatal depression | 25495 (1.0) | 657 (0.3) | 3333 (1.3) | 278 (0.6) |
| Pre-eclampsia | 5002 (0.2) | 591 (0.2) | 984 (0.4) | 158 (0.3) |

The observed 10-year CVD cumulative incidence was 3.18% (95% CI 3.07% to 3.29%) in Group C versus 2.09% (2.06% to 2.12%) in Group A. Figure 1B shows the observed 10-year cumulative incidence was generally higher in Group C than Group A across younger ages and in most ethnic groups, although some subgroup estimates were imprecise due to small numbers. At ages 40– 49, CVD incidence in Group C was 5.59% versus 1.68% in Group A in women and 6.07% versus 3.01% in men; at ages 50–59, 7.25% versus 3.59% and 8.50% versus 6.20%, respectively. Differences were also apparent but smaller below age 40.

Figure 2 illustrates the QRISK4-Lifetime risk for an example patient.

**Figure 2.**
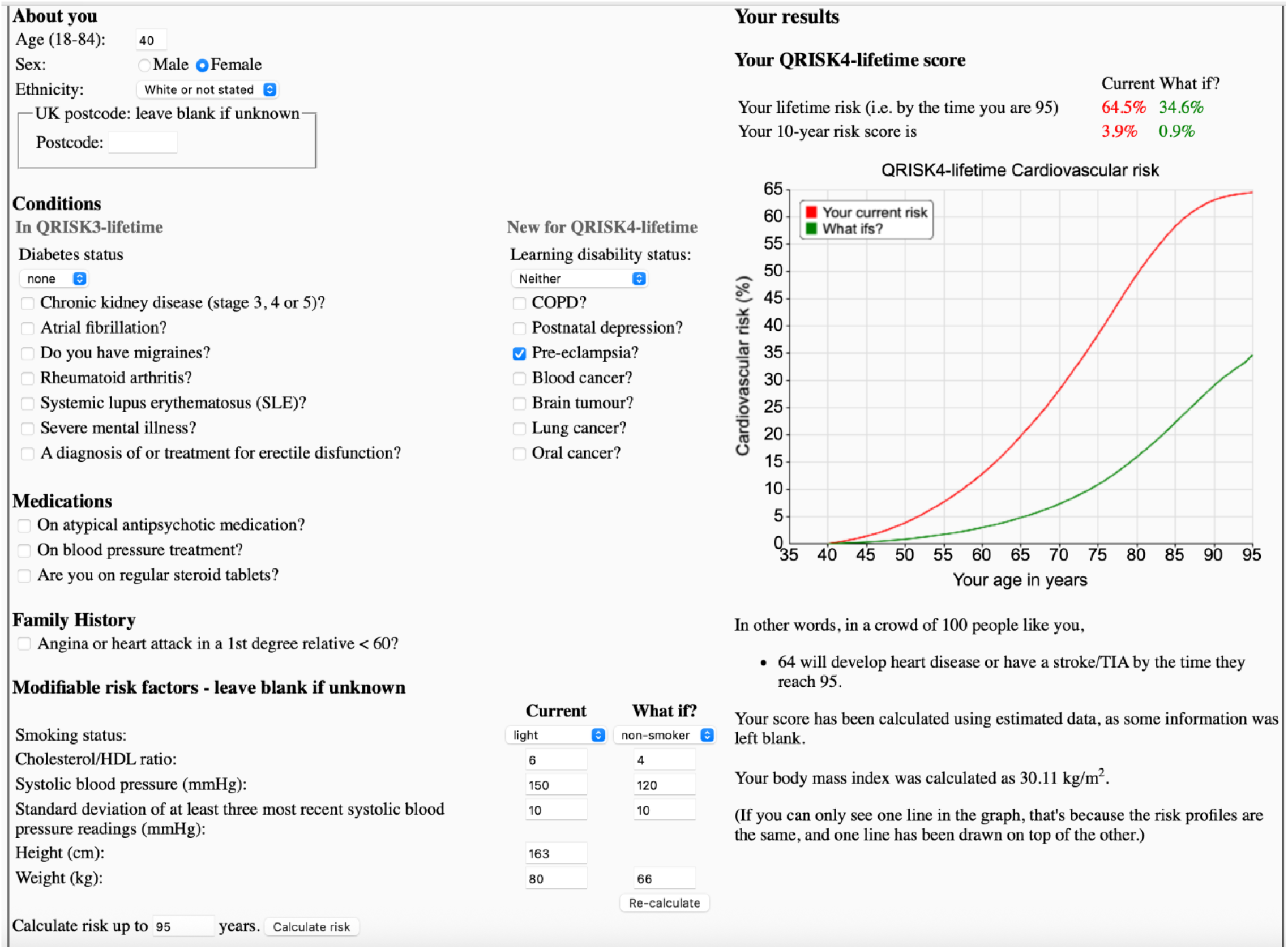
Web calculator illustration of a 40-year-old White women; with pre-eclampsia; light smoker; body mass index 30.1 kg/m^2^; cholesterol/HDL ratio of 6; systolic blood pressure 150 mmHg. The red line illustrates the predicted risk by age until the age of 95 which is 64.5%. Her 10-year risk is 3.9%. The green line shows a comparison with a 40-year-old White women; with pre-eclampsia who is a non-smoker; body mass index 25 kg/m^2^; cholesterol/HDL ratio of 4; systolic blood pressure 120 mmHg. The corresponding risk until the age of 95 is 34.6% with a 10-year risk of 0.9%.

## Discussion

We have developed and validated the QRISK4-Lifetime model which had similar discrimination and calibration values to QRISK4 in the separate validation cohort when evaluated at 10 years. However, the prediction horizon substantially changed who was selected as high-risk: almost three-quarters of patients meeting only the lifetime risk threshold were younger than 40. They fell below the UK’s specified 10-year risk threshold of 10%, although they might still receive preventive care if picked up opportunistically.

Participants meeting only the lifetime threshold were more likely to be South Asian, from deprived areas, be smokers, have hypertension, migraine or pre-eclampsia. They also had higher observed 10-year CVD incidence than those meeting neither threshold, particularly between ages 40-59. This was seen in both sexes and across ethnic groups. These descriptive comparisons support both short and longer-horizon risk assessment.

These findings illustrate how short-term 10-year assessment, in which age is a major determinant of absolute risk, gives a different perspective from assessment over the remaining lifetime. Lifetime estimates can support earlier discussions about smoking, other modifiable risk factors and prevention among people whose short-term risk remains low. This is consistent with NICE guidance supporting the use of QRISK3-Lifetime to inform risk discussions, particularly in younger adults or those with 10-year risk below 10%.^4^ European guidelines similarly support consideration of lifetime CVD risk in shared decision-making,^3^ while recent US guidance incorporates 30-year risk into selected lipid-lowering decisions.^1^ Canadian guidance recommends lifetime-risk counselling particularly focusing on adverse pregnancy outcomes.^2^ Whilst these approaches differ in their outcomes, populations and prediction horizons, our findings provide supporting evidence for longer-horizon risk assessment .

The illustrative lifetime risk threshold of 59.3% was chosen to match the number of individuals selected using the 10-year recommended threshold of 10% to enable a fair comparison. It does not represent an established threshold for treatment. It shows how lifetime assessment might complement short-term assessment: most high-risk patients (84.5%) selected by the 10-year threshold did not meet the lifetime threshold. Further research could establish whether communicating both results improves understanding, supports sustained risk-factor modification and leads to better clinical outcomes.

Study strengths include large, diverse routine-care cohorts, direct comparison of both models in the same validation cohort, and detailed performance assessment across age and ethnic groups. Lower discrimination within age-bands reflects more restricted variation in age and case mix. Limitations, including missing data and lack of formal adjudication of outcomes, mirror those in the original study.^8^ Lifetime estimates combine analyses across different ages and depend on assumptions about future hazards, risk factors and treatment. Importantly, the reclassification analysis is based on predicted risks and does not establish benefits of intervention in the newly selected group.

QRISK4-Lifetime provides an integrated framework for estimating CVD risk over two time horizons using the expanded QRISK4 predictor set within one tool. Its identification of a substantially younger high-risk group with a different ethnic and socioeconomic profile below the specified short-term threshold supports its potential role in preventive risk communication. Further research is needed to evaluate its clinical utility and cost-effectiveness and how best to communicate results.

## Data Availability

To guarantee the confidentiality of personal and health information only the authors have had access to the data during the study in accordance with the relevant license agreements. Access to the QResearch data is according to the information on the QResearch website (www.qresearch.org).

## Acknowledgements

We acknowledge the contribution of EMIS practices who contribute to QResearch and Queen Mary University of London for developing and supporting the QResearch database. We acknowledge the contribution of patient and lay representatives on the QResearch Scientific and Advisory Boards. This project involves data derived from patient-level information collected by the NHS, as part of their care and support. The Hospital Episode Statistics data, and civil registration mortality data are used by permission from NHS England who retain the copyright in that data. NHS England bears no responsibility for the analysis or interpretation of the data.

## Funding

This study was funded by Queen Mary University of London. There was no external funding.

## CODE AVAILABILITY

Clinical codes are published under a creative commons license, here https://www.qresearch.org/data/qcode-group-library/ with accompanying details in the Supplementary Table 1 of the original paper.^8^ Software implementing the QRISK4-Lifetime algorithm for research is available under an academic license.

## AUTHOR CONTRIBUTIONS STATEMENT

JHC initiated the study, development of the research question, undertook the literature review, data extraction, data manipulation and primary data analysis and wrote the first draft of the paper. CC contributed to the refinement of the research question, design, analysis, interpretation and drafting of the paper. JHC undertook final editing of the paper. Both authors commented critically on drafts of the manuscript and approved the final submission.

## COMPETING INTERESTS STATEMENT

The authors declare the following competing interests: JHC reports grants from National Institute for Health Research, and other research councils, during the conduct of the study. JHC is director of QResearch, a not-for-profit unit based at Queen Mary University of London. Until 2023, JHC had a 50% shareholding in ClinRisk Ltd, co-owning it with her husband, who was an executive director. In 2023, 100% of the share capital was donated to Endeavour Health Care Charitable Trust and the company renamed to Endeavour Predict Ltd. JHC is a consultant to Endeavour Predict Ltd. and her husband is a non-executive director. The company licenses software (including QRISK) both to the private sector and to NHS bodies or bodies that provide services to the NHS (through GP electronic health record providers, pharmacies, hospital providers and other NHS providers). CC reports receiving personal fees from ClinRisk Ltd until 2023.

## INCLUSION AND ETHICS

This study used effectively anonymised data from an electronic health care records database (QResearch) and hence participant consent was not required. The database covers a diverse population which is representative of the UK population. The QResearch ethics approval is given by the East Midlands-Derby Research Ethics Committee [Project OX330, ethics reference 23/EM/0166].

